# Reversal of increases in methadone distribution for opioid use disorder in the US during the COVID-19 pandemic

**DOI:** 10.1101/2022.03.09.22272154

**Authors:** Amy L. Kennalley, Jessica L. Fanelli, John A. Furst, Nicholas J. Mynarski, Margaret A. Jarvis, Stephanie D. Nichols, Kenneth L. McCall, Brian J. Piper

## Abstract

Opioid use disorder (OUD) is a major public health concern in the United States (US), resulting in high rates of overdose and other negative outcomes. Methadone, a treatment for OUD, has been shown to be effective in reducing the risk of overdose and improving overall health and quality of life. This study analyzed the distribution of methadone for the treatment of OUD across the US using data from the Drug Enforcement Administration’s Automated Reports and Consolidated Ordering System, Medicaid’s State Drug Utilization Data, and the US Census Bureau. Analysis revealed that methadone distribution for OUD has expanded significantly over the past decade, with an average state increase of +96.96% from 2010 to 2020, and there was a significant increase in overall distribution of methadone to opioid treatment programs (OTP) in the US from 2010 to 2020 (+61.00%) and from 2015 to 2020 (+26.22%). However, the distribution to OTPs did not significantly change from 2019 to 2021 (−5.15%). Furthermore, pronounced variation in methadone distribution among states were observed, with some states having no OTPs or Medicaid coverage. New policies are urgently needed to increase access to methadone treatment and address the opioid overdose crisis in the US.

## Introduction

The United States (US) Food and Drug Administration (FDA) has approved methadone, buprenorphine, and naltrexone as treatments for Opioid Use Disorder (OUD) [1], but there have been several policy changes impacting OUD care during the COVID-19 pandemic. Methadone is a long-acting synthetic opioid used to treat OUD via opioid treatment programs (OTP) [1-3]. Methadone take-home doses have been extended to 28 days for stable patients and 14 days for less stable patients, drug screening requirements have been relaxed, and telemedicine has been expanded for established patients [4, 5]. However, unlike for buprenorphine, starting methadone treatment required an in-person visit. Prior to the pandemic, OUD methadone treatment also required the co-administration of counseling, however, these counseling requirements have also been relaxed as a result of the COVID-19 pandemic [5-7].

Methadone treatment, though considered a gold standard treatment for OUD, is not without serious adverse effects similar to other opioids: respiratory depression and cardiotoxicity [3, 8, 9]. An alternative standard treatment to methadone, buprenorphine, has also raised safety concerns as well with rising respiratory depression fatalities after oral doses in both adult and pediatric patients [10]. It is important to contextualize these concerns with the extremely high risk of respiratory depression associated with ongoing nonprescribed IV opioid use, a symptom of under or untreated OUD. In regions where it is available, methadone is frequently used to treat the most severely ill OUD patients since it is a more potent opioid receptor activator than buprenorphine. This strong impact necessitates expert handling, especially in the early stages of treatment [11]. In addition to its adverse effects, buprenorphine was widely but inequitably unavailable from 2004 to 2015 due to factors such as race and socioeconomic status, finding that Black patients had a lower probability of receiving a prescription [12]. To further bolster the use for methadone for OUD, treatment with methadone was superior to buprenorphine in reducing criminal activity, HIV infection, hepatitis, and overall mortality [2,13-16]. Additionally, a Cochrane Review concluded that methadone was superior to buprenorphine in retaining patients in treatment (Supplemental Table 1) [17]. However, there is a nationwide lack of appropriate treatment with all potential medications for persons with OUD [18, 19].

There have been over one-million drug overdoses in the US since the start of the opioid epidemic [20]. The COVID-19 pandemic has placed tremendous stress on the healthcare system including the access to providers and availability of OUD treatments. Between 2019 and 2020, there was a +48.8% increase in overdose mortality among Black people, compared to +26.3% among White people [21]. Moreover, from April 2020 to April 2021, the number of drug overdoses in the US exceeded one-hundred thousand, a +28.5% increase over the previous year [22]. With a +60% rise in overdoses compared to the previous year, May 2020 became the deadliest month on record [23]. A cross-sectional study, conducted from May to June 2020 in the US and Canada, found that new patients wishing to initiate methadone treatment were faced with a barrier in 20% of clinics [24]. Similarly, prior to the pandemic, both methadone and buprenorphine-based OTPs were found to be effective in US jails and prisons. However, following the pandemic, some of these OTPs have been discontinued while others have been expanded [25-27].

This study obtained data from the Drug Enforcement Administration’s (DEA) Automated Reports and Consolidated Ordering System (ARCOS), a federal program established by the 1970 Controlled Substances Act, to monitor the distribution of DEA controlled substances from various sources including retail pharmacies, hospitals, practitioners, teaching institutions, mid-level practitioners, and OTPs. Previous pharmacoepidemiologic studies have also utilized the ARCOS database [28-32]. It is important to note that ARCOS does not provide information on the number of patients receiving methadone. This caveat is important because it prevents an accurate representation of the amount of methadone used for each OUD patient. However, it is worth mentioning that federally funded OTPs do keep track of the number of patients receiving treatment, which could be a useful resource in understanding the true scale of the OUD epidemic in the US and the effectiveness of treatment efforts. The number of patients receiving methadone for OUD decreased by almost one-quarter (−23.7%) from 2019 (408,550) to 2020 (311,531) [33].

In addition to ARCOS, Medicaid’s State Drug Utilization Data (SDUD) database was used in this study [34]. Medicaid is a program at the federal and state level which functions to aid in covering healthcare costs for patients with limited resources [35]. Medicaid.gov publishes all prescription drugs covered by Medicaid every year for all 50 states and the District of Colombia (DC) in the SDUD database. The State Health Official Letter, released on December 30, 2020, states that the SUPPORT Act of 2018 mandates the inclusion of Medicaid coverage for medication-assisted treatment for eligible OUD patients. Subsequently, the Continuing Appropriations Act of 2021, which added to the SUPPORT Act, requires rebates on methadone and other OUD drugs starting from October 1, 2020 to September 30, 2025. [36, 37]. The use of both ARCOS and SDUD databases provide a comprehensive picture of the distribution and utilization of methadone for the OUD treatment over the past decade. Together, it is critical to examine the changes in methadone distribution during the COVID-19 pandemic and determine whether there are national or regional barriers to access this evidence-based pharmacotherapy.

## Material and Methods

### Procedures

The quantities of methadone distributed (in grams) were obtained from the ARCOS yearly drug summary reports for 2010, 2015, 2019, 2020 and 2021. Methadone distributed to OTPs was classified as an OUD treatment. The number of OTPs per state were also obtained from ARCOS for the years 2010, 2015, 2019, 2020 and 2021. The state population estimates, including DC, were derived from the US Census Bureau’s Annual Estimates of the Resident Population for the US [38]. The US Territories were examined elsewhere and were not included in Figures 1-5 [39]. Data was collected for methadone covered by Medicaid in 2020 for all 50 states and DC using a filtered download from the SDUD database [34]. National Drug Codes of formulations that are primarily used for OUD are provided in the Supplemental Table 2. The number of methadone prescriptions per state was divided by the number of Medicaid enrollees per state in 2020. Three states—Virginia, Montana, and Iowa (Supplemental Figure 1 also shows 2019 and 2021) were excluded from the results due to being outliers (10-112K prescriptions/100K enrollees) which presumably reflected a Medicaid data error (mean 475.4 prescriptions/100K enrollees). This study was approved by the institutional review boards of the University of New England and Geisinger.

**Figure 1.**
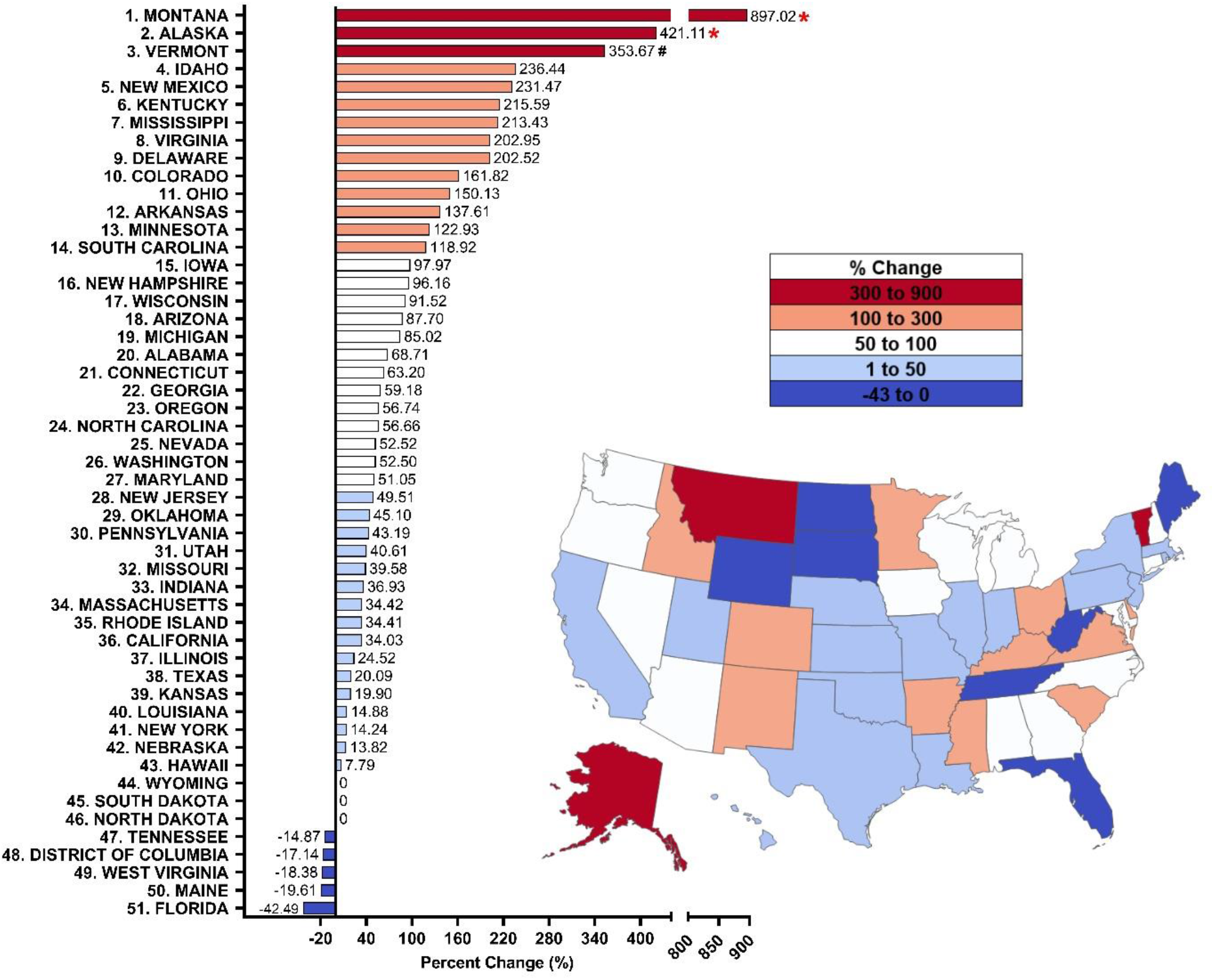
Percent change from 2010 to 2020 in methadone distribution as reported by the Drug Enforcement Administration’s Automated Reports and Consolidated Ordering System for Opioid Use Disorder. Percent change between 1.5 SDs and 1.959 SDs from the mean (+96.96%, SD = 146.64%), indicated with a #. Percent change > ±1.96 SD from the mean was considered significant (*P < 0.05).

### Data Analysis

The percent change in methadone distribution for OUD was compared between states for time spans of ten years, five years and one year respectively. The distribution of methadone, corrected for population, was compared between 2010 and 2020, 2010 and 2021, 2015 and 2020, 2015 and 2021, 2019 and 2020, 2019 and 2021, and 2020 and 2021 were considered significant if the paired t-test indicated P < 0.05. Any state with percent changes greater than ±1.96 SDs from the national average percent change was considered statistically significant. The number of OTPs per one million persons per state between years—2010 and 2015, 2010 and 2020, 2010 and 2021, 2015 and 2020, 2015 and 2021, 2020 and 2021—were considered significant if the t-test indicated P < 0.05. Heatmaps created using JMP version 16.2.0. Figures and data analysis completed using Microsoft Excel and GraphPad Prism version 9.4.0.

## Results

### ARCOS

Overall, the total volume of methadone distributed to OTPs in the US increased over the last decade. The amount distributed to OTPs was 8.62 metric tons in 2010, 10.88 tons in 2015 (+26.3%), and 13.03 tons in 2020 (+19.7%), reflecting an increase in distribution to the majority of states. Specifically, the national distribution of methadone for OUD from 2010 to 2020 increased by +96.96% (SD=146.64) on average, with forty-three states showing an increase, five states a decrease, and three states showing no change. A t-test revealed a significant increase (+61.8%) from 2010 to 2019 (P < 0.0001) and by +61.0% from 2010 to 2020 (P < 0.0001). From 2010 to 2020, there was a large (> 1.5 SDs) increase in Vermont (+353.67%), and significant elevations (> 1.96 SDs, P < 0.05) in Alaska (+421.11%) and Montana (+897.02%) relative to the average (Figure 1).

Examination of 2015 relative to 2020 revealed the national average distribution for OUD increased by +26.22% (SD = 50.38%), with thirty-eight states increasing but eleven states decreasing. A t-test revealed a significant (P < 0.0001) increase. There were significant increases (P < 0.05) in Alaska (+135.34%) and Mississippi (+311.48%) relative to the national mean (Figure 2).

**Figure 2.**
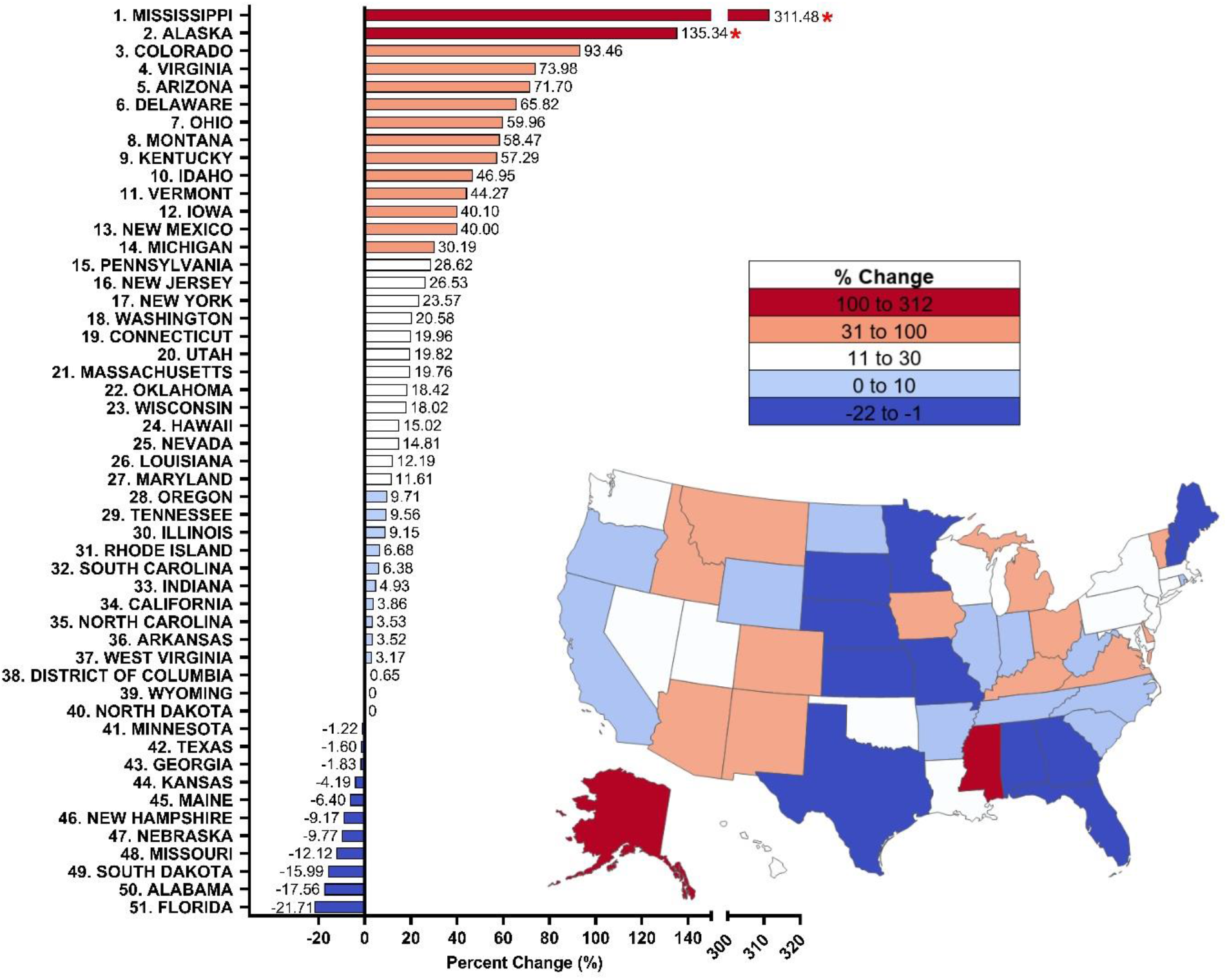
Percent change from 2015 to 2020 in methadone distribution as reported by the Drug Enforcement Administration’s Automated Reports and Consolidated Ordering System for Opioid Use Disorder. Percent change between 1.5 SDs and 1.959 SDs from the mean (+26.22%, SD = 50.38%), indicated with a #. Percent change > ±1.96 SD from the mean was considered significant (*P < 0.05).

However, from 2019 to 2020 (i.e., pre-to post-COVID-19 pandemic), the distribution of methadone was stable (−0.091%, SD = 10.81%). Slightly over half (twenty-eight) of states showed an increase and twenty-two states exhibited a decrease. No significant (P = 0.76) differences were found between 2019 and 2020. Examination of specific states revealed an increase in Kentucky (+18.68%) and a significant increase (P < 0.05) in Ohio (+26.02%) relative to the national mean. In contrast, there were appreciable decreases in Nebraska (−16.6%), South Dakota (−17.27%), and Mississippi (−20.53%), and significant decreases (P < 0.05) in Alabama (−21.96%), New Hampshire (−24.13%) and Florida (−28.97%) relative to the national mean (Figure 3).

**Figure 3.**
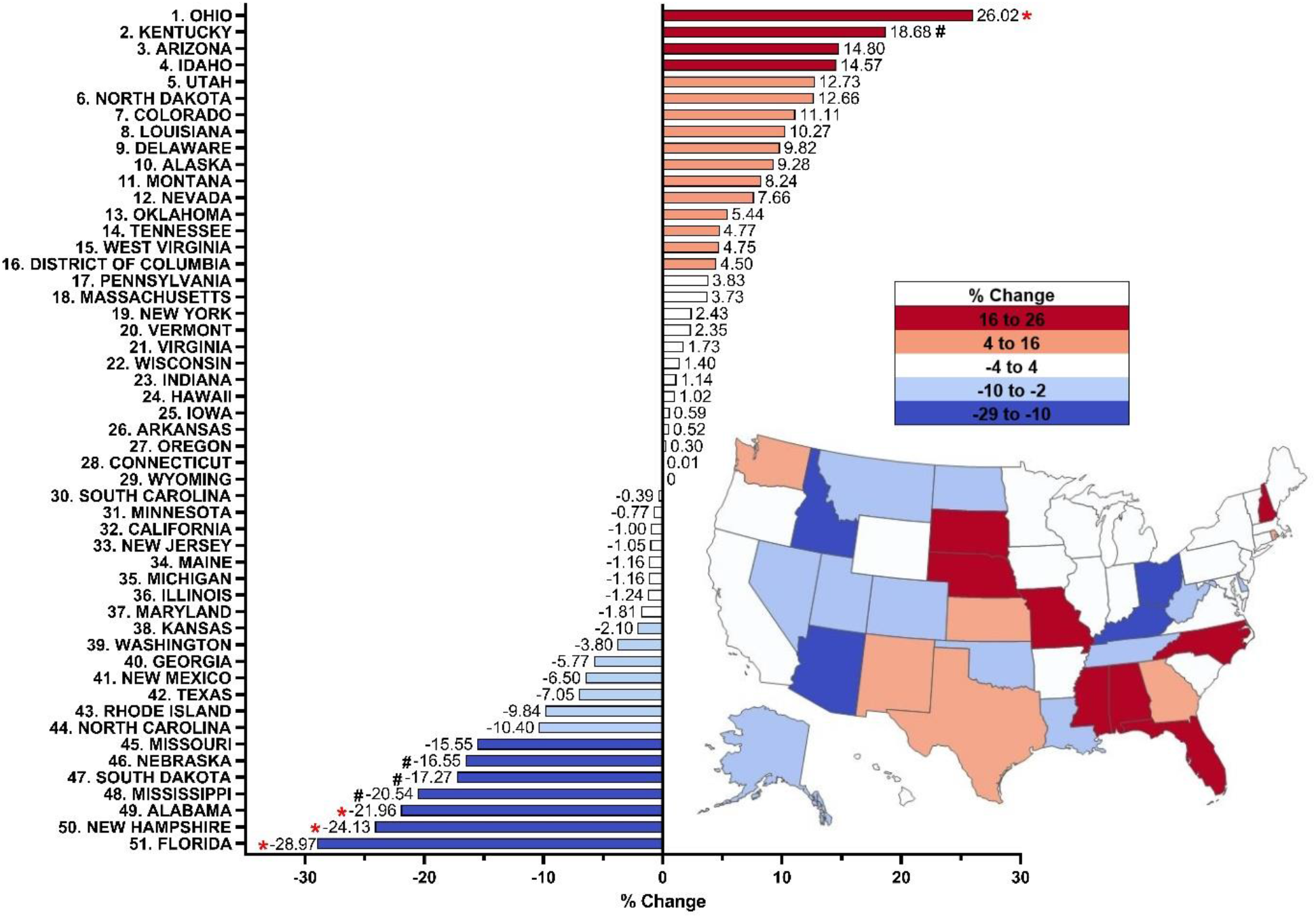
Percent change from 2019 to 2020 in methadone distribution as reported by the Drug Enforcement Administration’s Automated Reports and Consolidated Ordering System for Opioid Use Disorder. Percent change between 1.5 SDs and 1.959 SDs from the mean (−0.09%, SD 10.81), indicated with a #. Percent change > ±1.96 SD from the mean was considered significant (*P < 0.05).

Looking at 2019 to 2021, a wider pre-to post COVID-19 pandemic timeline, the distribution of methadone from OTP showed a decline (−5.15%, SD = 19.14%). Eighteen states showed an increase while thirty-one states showed a decrease. No significant (P = 0.065) differences were found between 2019 and 2021. Ohio (+47.53%) had a significant increase (P < 0.05) relative to the national mean. In contrast, there was an appreciable decrease in Mississippi (−42.53%), and significant decreases (P < 0.05) in Alabama (−44.27%), Nebraska (−47.96), New Hampshire (−52.99%) and Florida (−54.61%) relative to the national mean (Figure 4). However, distribution in 2021 was - 5.69% lower than 2019 and - 5.71% lower than 2020 (Supplemental Figure 2A).

**Figure 4.**
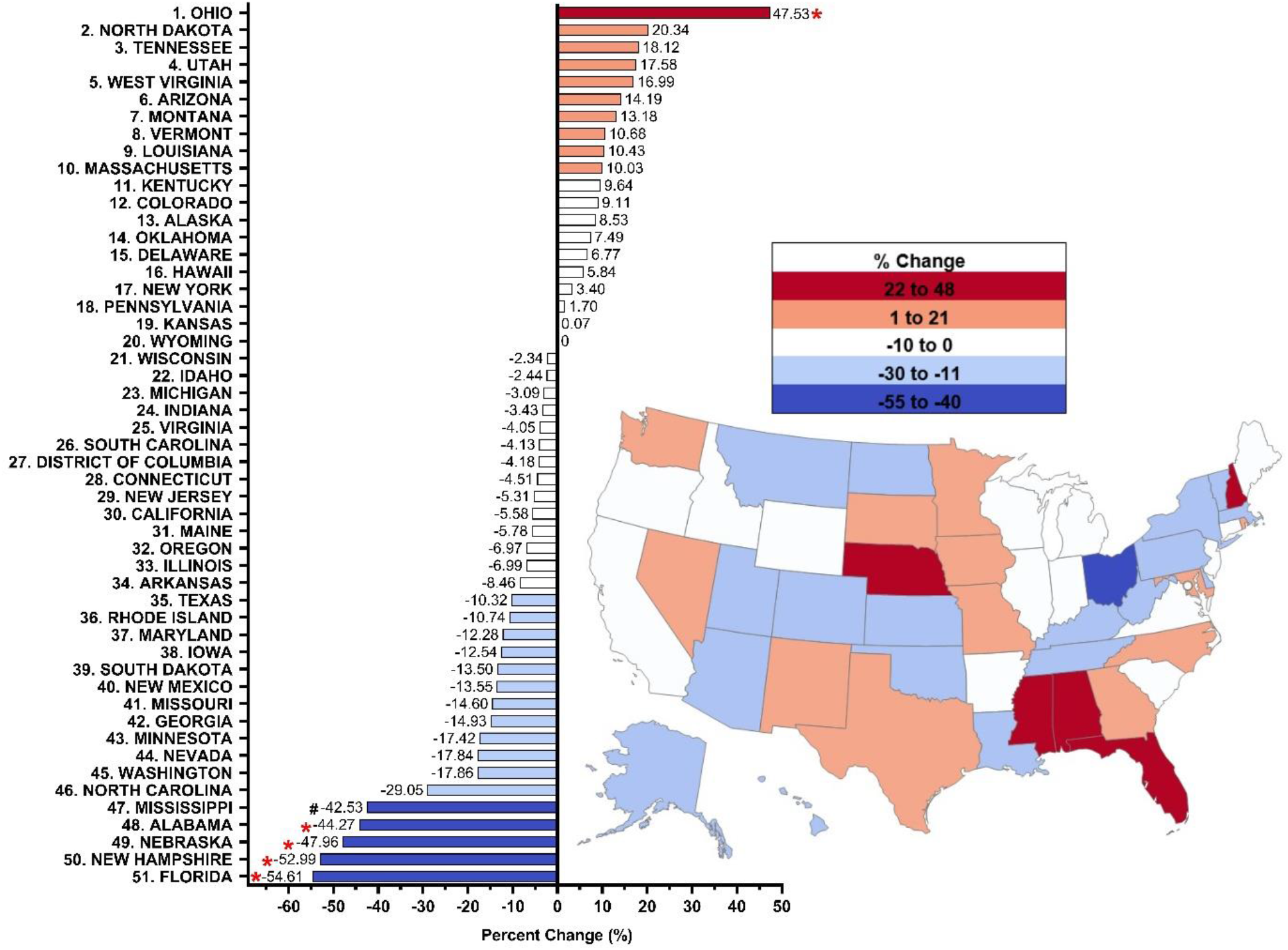
Percent change from 2019 to 2021 in methadone distribution as reported by the Drug Enforcement Administration’s Automated Reports and Consolidated Ordering System for Opioid Use Disorder. Percent change between 1.5 SDs and 1.959 SDs from the mean (−5.15%, SD 19.14), indicated with a #. Percent change > ±1.96 SD from the mean was considered significant (*P < 0.05).

The average methadone distribution in the US for OUD was 43.75 mg/person (SD = 35.01) in 2021. There was significantly elevated methadone distributed in Rhode Island (155.13 mg/person), Delaware (147.27 mg/person), Connecticut (126.66 mg/person), and Vermont (125.06 mg/person) relative to the national mean (43.75, P < 0.05).

The total number of OTPs distributing methadone in the US increased +18.6% from 2010 (1,139) to 2015 (1,351) and peaked in 2021 (1,738, Supplemental Figure 2B). The number of pharmacies distributing buprenorphine (49,041) was 43.1-fold greater than OTPs distributing methadone in 2010. However, this ratio decreased to 33.3-fold in 2021. The number of OTPs per 1 million persons per state significantly increased (P < 0.0001) from 2010 to 2021. A t-test indicated that 2010 to 2015 (P < 0.0001), 2010 to 2020 (P < 0.0001), 2010 to 2021 (P < 0.0001), 2015 to 2020 (P < 0.0001), and 2015 to 2021 (P < 0.0001) were each significantly different. The number of OTPs per 1 million persons per state in 2019 relative to 2021 (P = 0.0652), and 2020 relative to 2021 (P = 0.1741) did not significantly increase. Twenty states had fewer OTPs in 2021 relative to 2010, 2015, or 2020 (Maryland, Vermont, New Mexico, Maine, Georgia, North Carolina, New York, New Hampshire, Utah, Oklahoma, South Carolina, Montana, Washington, Alabama, Texas, Minnesota, Florida, South Dakota, Nebraska, and Mississippi). One state (Wyoming) did not have a single OTP (Figure 5, Supplemental Table 3).

**Figure 5.**
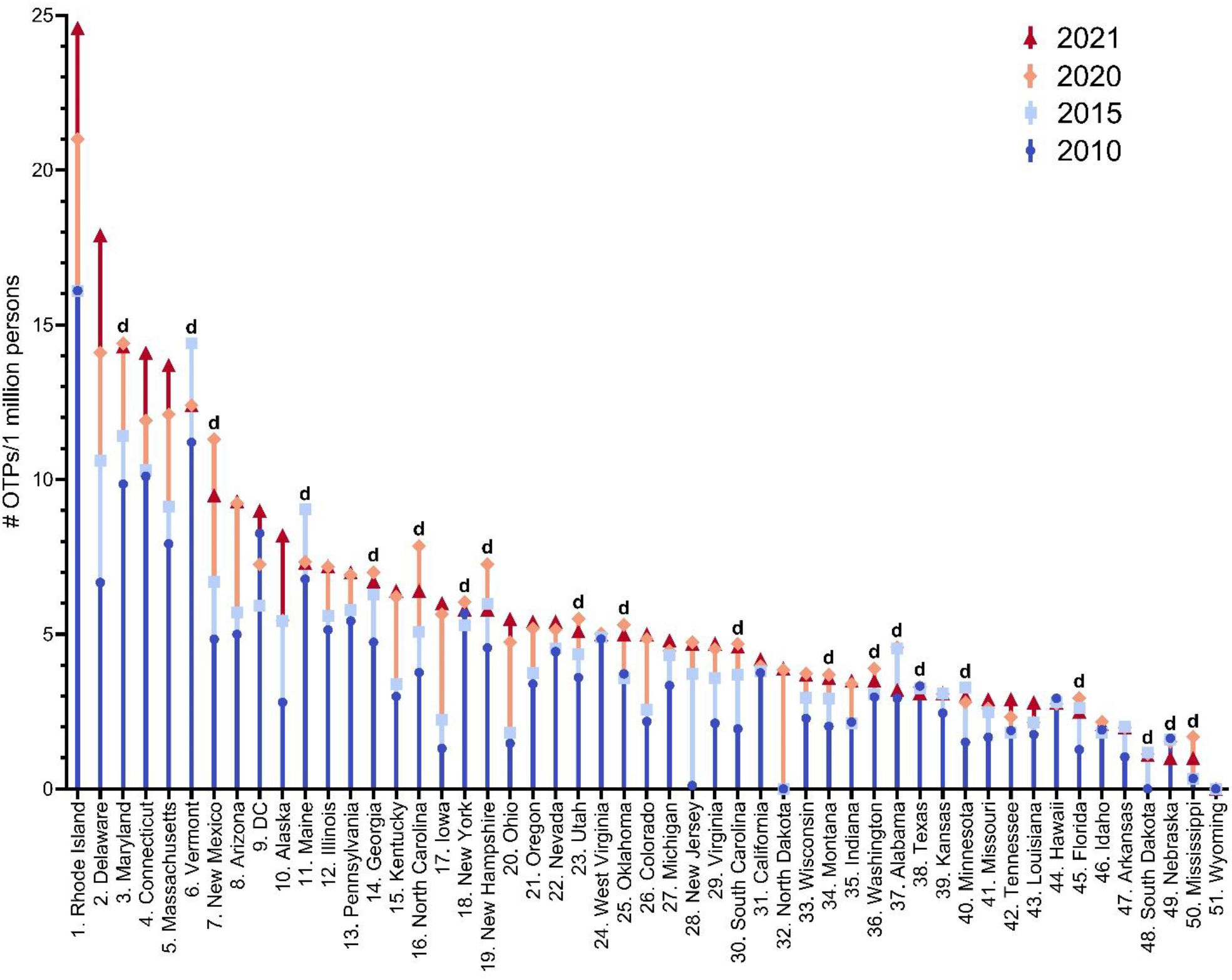
Number of opioid treatment programs per 1 million persons per state all significantly (P < 0.0001) different from 2010, 2015, 2020 and 2021. The twenty states that decreased in 2021 relative to 2010, 2015 or 2020 are indicated with a “d”. DC: District of Columbia.

### Medicaid

The SDUD database showed considerable variation in methadone prescribing between states and regions for patients covered under Medicaid (mean = 475.39, SD = 1097.78 with four states having values of 0). The top four states (Wisconsin, Tennessee, Oregon, and Vermont) accounted for 64.03% of all methadone covered by Medicaid in 2020 (Supplemental Figure 3).

## Discussion

The key finding of this study was that the pronounced and significant increase in methadone distribution to OTPs for OUD over the past decade has reversed with a non-significant decrease (−0.09%) from 2019 to 2020, and a decline (−5.15%) from 2019 to 2021. There were also significant increases in the number of OTPs per one million persons per state over the past decade, but no significant increases over the COVID-19 pandemic period from 2019 compared to 2021 [28]. These findings point to the necessity for more OTPs to combat the escalating OUD problem. Twenty states showed a reduction in OTPs from 2010 to 2021 which could be due to funding issues or policies for OTPs in these states [40].

Looking at how the COVID-19 pandemic affected OUD treatment from 2019 to 2021, the findings of this study show methadone distribution to OTPs did not increase significantly. However, a subtle but statistically significant increase (+5.0%) was observed in the number of poison control reports of intentional methadone exposure during this period [41]. In addition, the National Center for Health Statistics (NCHS) of the Centers for Disease Control and Prevention (CDC) reported that drug overdose mortality increased by +31% between 2019 and 2020. Nonetheless, the rate of methadone overdose deaths remained low with no significant increase in this report, suggesting no change in methadone misuse during the COVID-19 pandemic [42]. However, others have reached the opposite conclusion [9]. Since OTPs were not distributing additional methadone, on an MME level, during the ongoing COVID-19 pandemic, this emphasizes the need for methadone treatment, as well as the necessity for safe storage and expanded availability to prevent opioid overdoses. Individuals who overdose may be prime candidates for methadone treatment. It may be worth exploring options such as allowing for limited take home access at community pharmacies after a period of OUD stability, which is currently prohibited by law outside of an OTP [43]. Another uncommonly employed solution is to provide travelling methadone treatment on a daily route which can allow observed administration and decrease need for transportation to a further location [44].

As of 2018, forty-one states had methadone covered under Medicaid [45]. Despite methadone being an evidence-based treatment of OUD and pain management, twelve states located predominantly in the South and Midwest opted to not expand Medicaid, leaving patients in these areas vulnerable to inaccessible treatment for OUD and for pain [46, 47]. States where methadone was not covered included: Arkansas, Idaho, Kansas, Kentucky, Louisiana, Nebraska, North Dakota, South Carolina, Tennessee, and Wyoming [45]. Although over four fifths of states can, in theory, treat Medicaid patients with methadone for OUD, we found that only four states accounted for the preponderance (64.03%) of prescriptions. This pronounced disparity in methadone coverage under Medicaid between states indicates that patients in certain states and regions have better access to this evidence-based treatment for OUD than others [2]. Moving forward, improvements in access to methadone prescriptions and clinics for Medicaid patients need to be expanded in order to equalize treatment options for this large, but vulnerable, population. Having coverage for methadone under Medicaid in all fifty states will help combat the escalating opioid epidemic by expanding treatment of substance use disorder (SUD) and OUD [45]. There is also a need for grant funding request for proposals to specifically focus on programs aimed at increasing access to methadone, particularly in rural areas.

This study suggests the importance of policy change as it is apparent there is a pressing need for additional access to OUD therapy, specifically methadone treatment, in the US. Over 400,000 people in the US received methadone from an OTP pre-pandemic, 2019, with over 90% located in urban areas, making it challenging for rural patients to make the daily trip to receive their medication [48, 49]. SAMHSA released guidelines in March 2020 allowing the regulatory authorities of all states to request blanket exceptions to allow OTP patients to take home doses of methadone and buprenorphine [50]. These guidelines were extended in November 2021. However, the number of patients receiving methadone declined in the first year of the COVID-19 pandemic by one-quarter [33]. Many providers and public health researchers are calling for these take home dosage rules to be continued post-pandemic [51, 52], although others that evaluated the number of overdoses involving methadone are more cautious [9, 53]. The number of pharmacies which dispensed buprenorphine was 34-fold greater in 2021 than the number of OTPs that dispensed methadone. It is currently curious that there are more restrictions in the US for prescribing methadone than for other Schedule II substances like fentanyl or oxycodone. Overcoming the “Not in my Backyard” (NIMBY) stigma surrounding OTPs is not a trivial undertaking [39]. It will require a paradigm shift to allow supervised administration in pharmacies, primary care offices, and mobile units as are common in other Western nations [13, 24, 54]. Expanding the role of specialty trained Board Certified Psychiatric Pharmacists by loosening the medication for opioid use disorder (MOUD) regulations to allow already prescribing pharmacists to obtain an X-DEA number, can immediately increase access to MOUD [55, 56]. According to the Consolidated Appropriations Act, 2023, which was passed on December 29, 2022, medical providers with a current DEA registration number are now able to prescribe buprenorphine for OUD without needing an X-DEA waiver if state law permits it [57]. Overall, this is important because it aims to remove existing barriers to OUD treatment and increase access to care for individuals who need it.

The strengths of this report include novel and timely data from ARCOS which is comprehensive for both distribution and number of distributors. This investigation extends upon earlier research both pre [4] and post COVID-19 pandemic [31, 32]. Potential caveats and limitations stem from the fact that methadone distribution is reported in ARCOS by weight rather than prescriptions per individual at an OTP. Importantly, other pharmacoepidemiological research that compared another Schedule II substance from ARCOS to a state Prescription Drug Monitoring Program identified a high correspondence (r = +.985) [30]. The inclusion of Medicaid data also, at least partially, offsets this concern. The paucity of methadone OTPs across rural states like Wyoming (253k km^2^), South Dakota (200k km^2^), and Nebraska (200 km^2^) is an important finding. However, prior research found that another rural state, Maine, which ranked tenth in the US, had three-OTPs in a single thirty-thousand-person city (Bangor) and none in other areas in northern Maine [49]. The non-homogenous distribution within states should also be a concern for policy makers. Providing funding opportunities to train Addiction Medicine Fellows who will focus on rural care post-graduation can result in a pipeline of addiction medicine leaders to many areas of the US which are most in need. Future research should also be focused on determining which patient subgroups were most impacted by the reversal in methadone distribution. As the number of pharmacies nationally distributing buprenorphine decreased (−4.7%) from 2019 to 2021, further research should evaluate if both methadone and buprenorphine continue to be underutilized in the post-COVID-19 pandemic period [58] (Supplemental Table 1).

## Conclusions

In conclusion, this study identified a reversal over the past-decade in the distribution of methadone for OUD in the US. The pronounced OTP geographical disparities, particularly in rural and western states, is a long-standing concern. The many policy accommodations to COVID-19 may present an important opportunity to determine which factors most impact the accessibility, adherence, safety, and efficacy of methadone.

## Data Availability

All data produced are available online at the DEA's Automated Reports and Consolidated Ordering System and Medicaid's State Drug Utilization Data.

https://www.deadiversion.usdoj.gov/arcos/retail_drug_summary/index.html

https://nam10.safelinks.protection.outlook.com/?url=https%3A%2F%2Fwww.medicaid.gov%2Fmedicaid%2Fprescription-drugs%2Fstate-drug-utilization-data%2Findex.html&data=05%7C01%7CAKennalley%40som.geisinger.edu%7Cbf1fa1b23daa469ac51f08daaec32aad%7Cc048947985ac4b82bbad54c03fc21678%7C0%7C0%7C638014450845928960%7CUnknown%7CTWFpbGZsb3d8eyJWIjoiMC4wLjAwMDAiLCJQIjoiV2luMzIiLCJBTiI6Ik1haWwiLCJXVCI6Mn0%3D%7C3000%7C%7C%7C&sdata=xZLVJRHiS%2BoukxF0vELEvnUhD2aSrTwo3sz5u%2BV7fig%3D&reserved=0

---

**Supplemental Figure 1.**
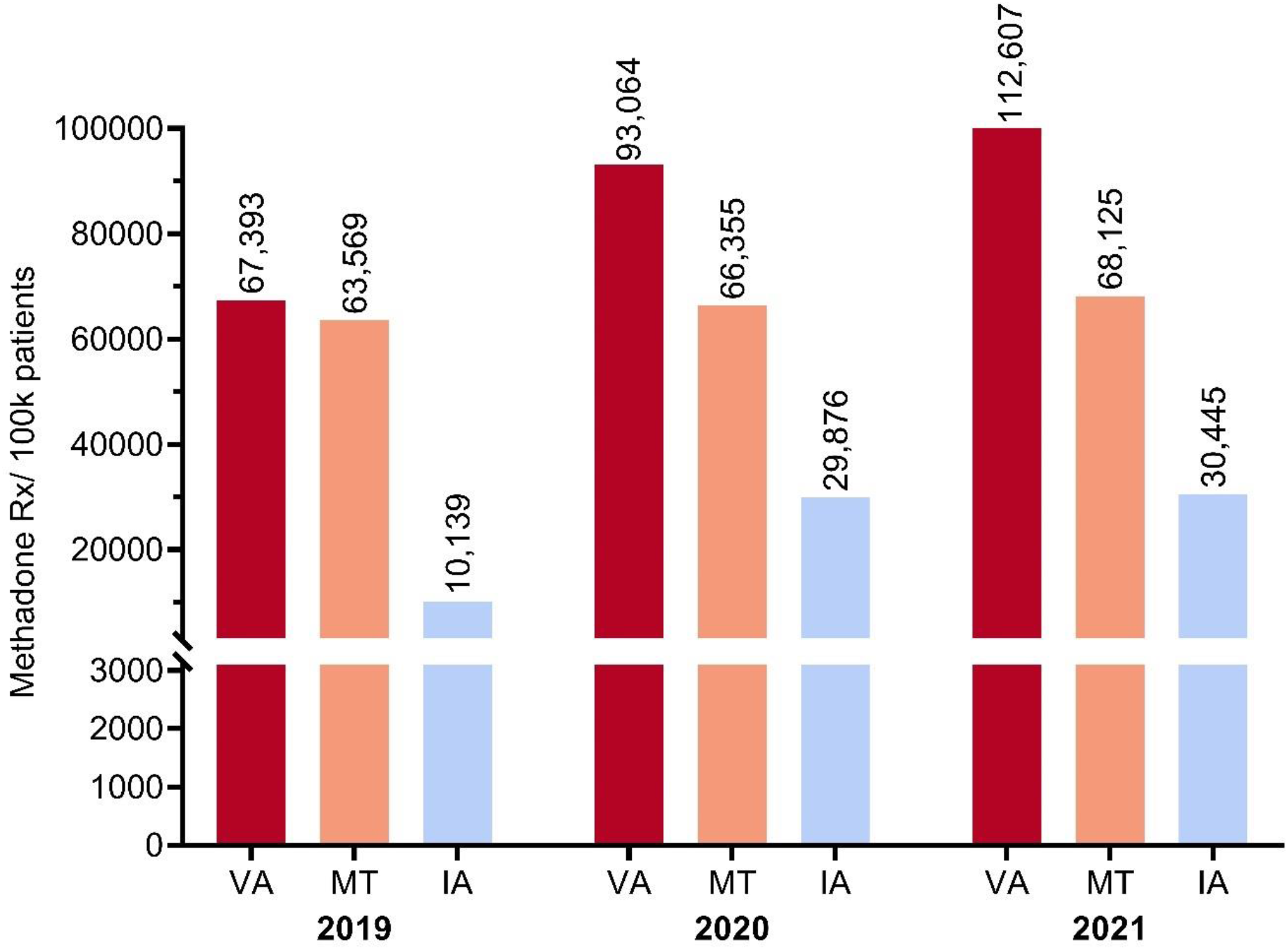
Methadone prescriptions per 100K Medicaid patients, for top three US states Virginia, Montana and Iowa, for the years 2019, 2020, and 2021.

**Supplemental Figure 2.**
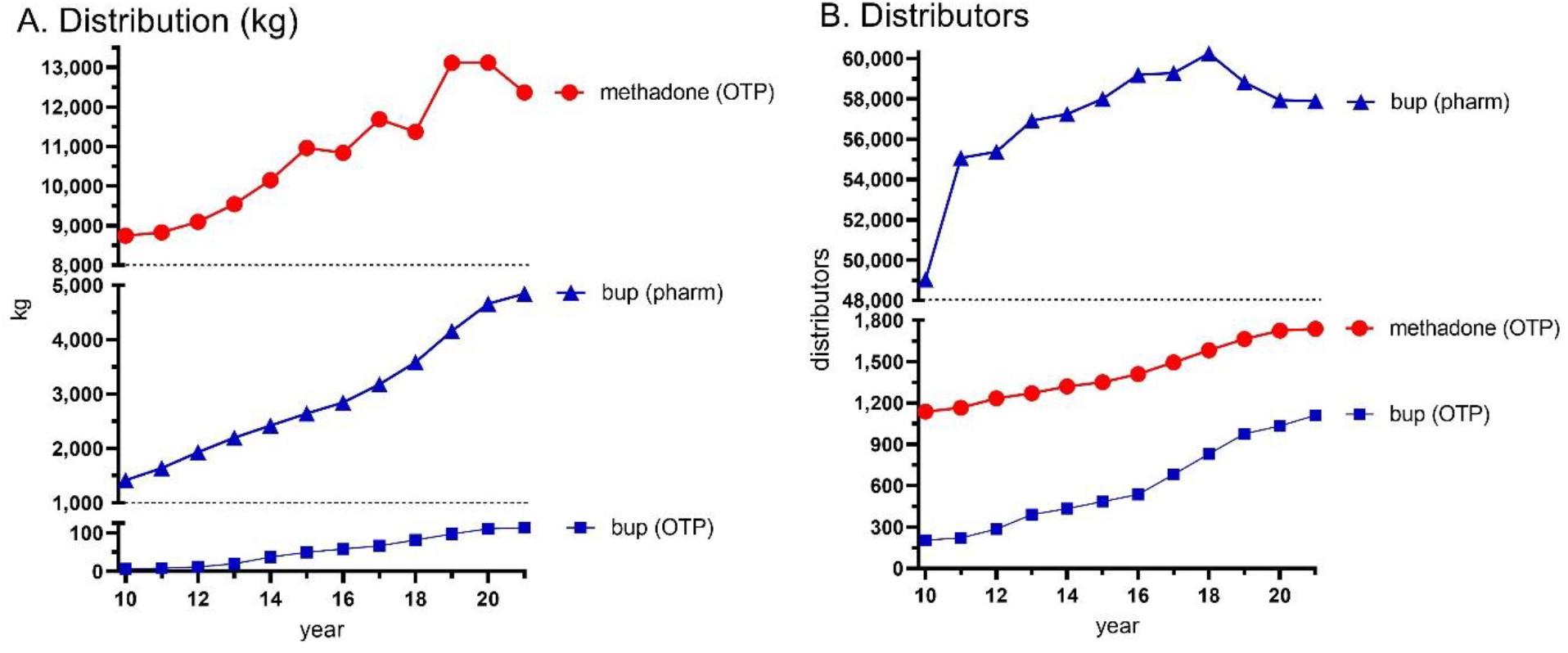
Distribution in kg (A) and number of buyers and registrants (B) in Opioid Treatment Programs (OTP) and pharmacies (pharm, buprenorphine only) for methadone and buprenorphine as reported to the United States Drug Enforcement Administration’s Automated Reports and Consolidated Orders system for the fifty states, Washington DC, and the US Territories. Methadone in 2021 (12.4 metric tons) was 5.69% lower than 2019 (13.1 metric tons) and 5.71% below 2020 (13.1 metric tons). Methadone by weight was 2.49-fold greater than buprenorphine from pharmacies and OTPs in 2021. The number of pharmacies distributing buprenorphine was 33.3 to 47.3-fold greater than the number of OTPs distributing methadone from 2010 to 2021.

**Supplemental Figure 3.**
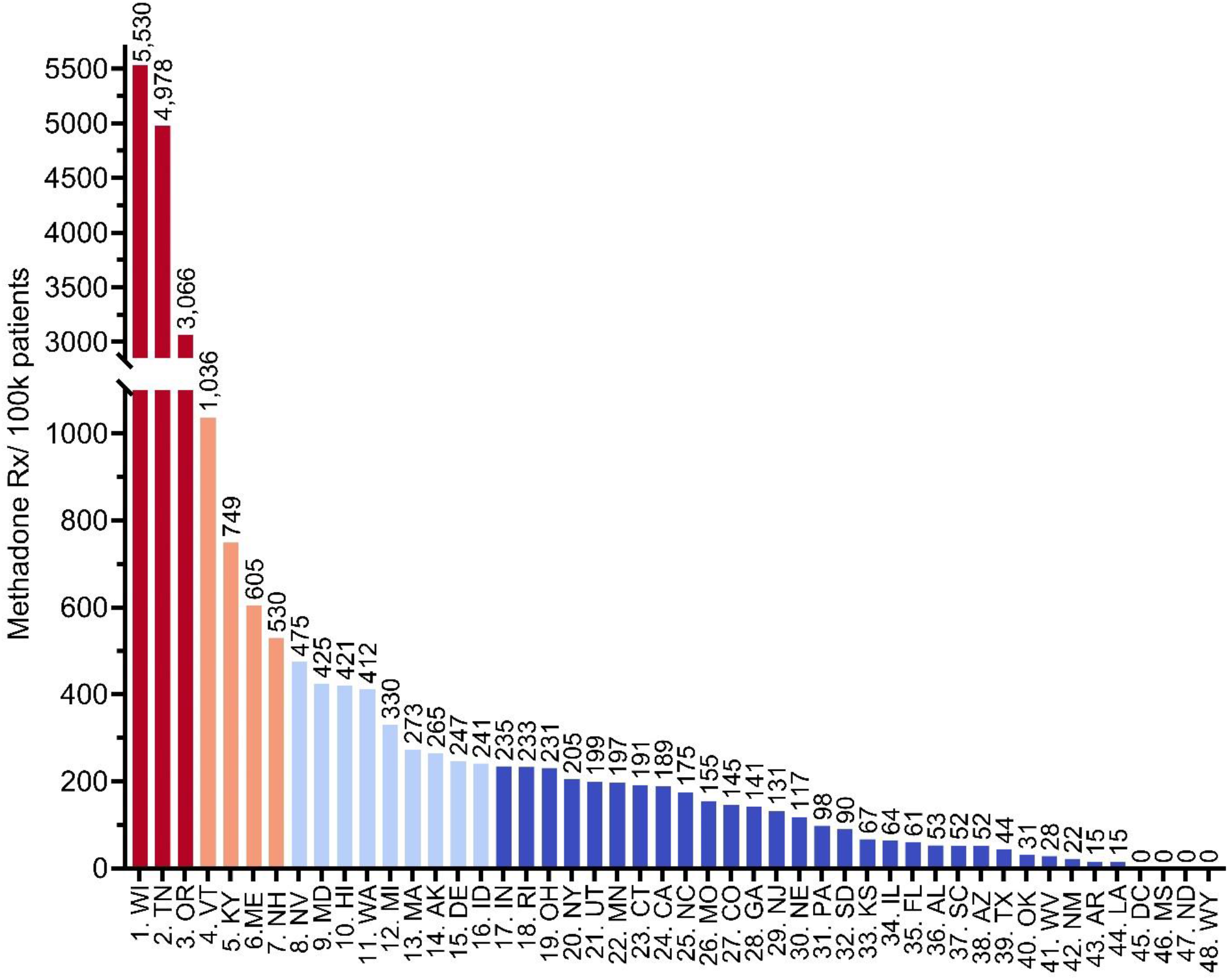
Methadone prescriptions, per 100K Medicaid patients, for 48 US states, and Washington DC.

**Supplemental Table 1.**
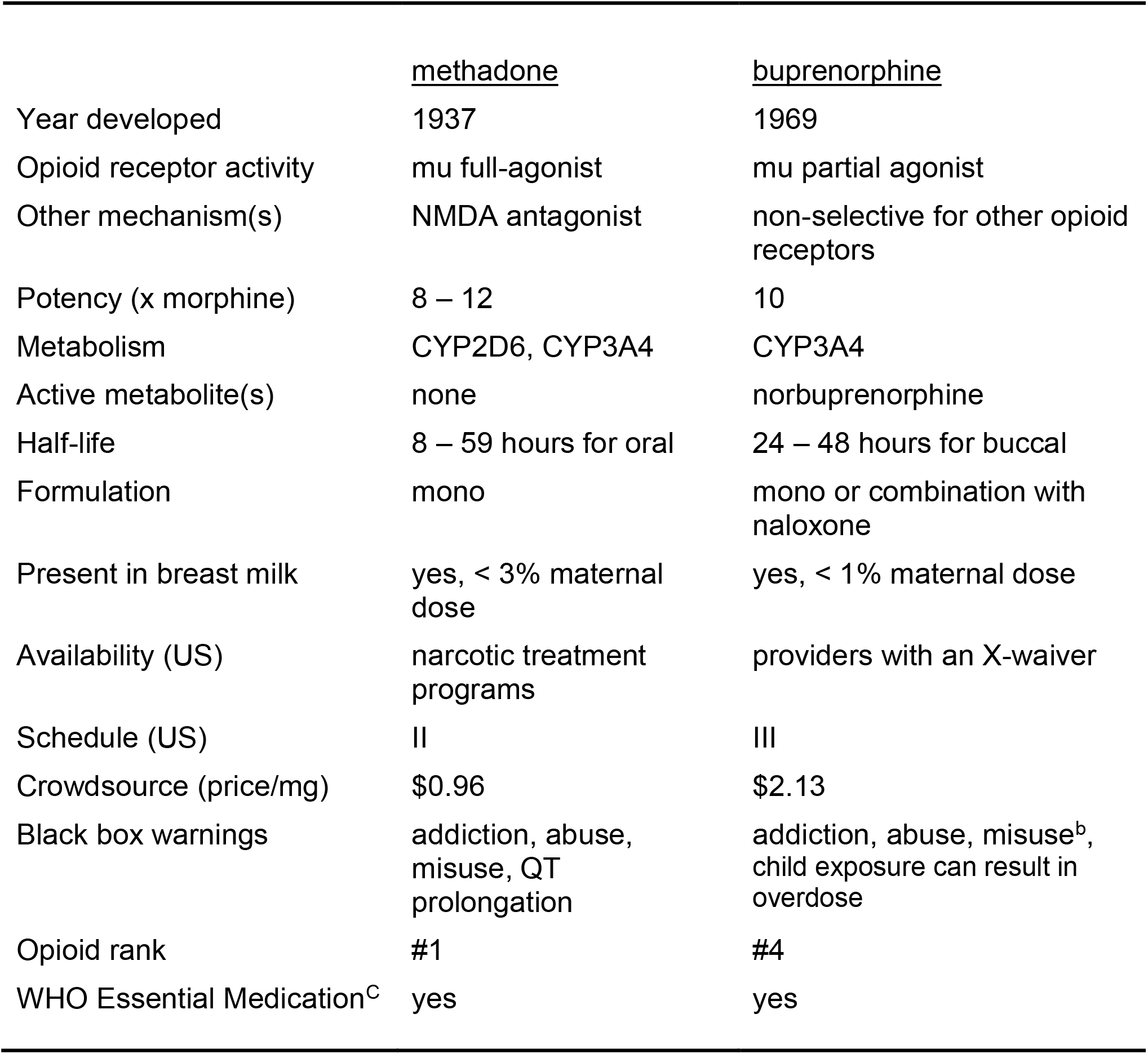
Comparison of the pharmacological and therapeutic properties of methadone and buprenorphine. ^b^buccal formulation, ^C^complimentary

**Supplemental Table 2.** Medicaid’s State Drug Utilization Data reported National Drug Codes (NDC) for methadone prescribed for opioid use disorder.

| **NDC** |
| --- |
| 54039168 |
| 54039268 |
| 54355344 |
| 54355563 |
| 54421825 |
| 54421925 |
| 54453825 |
| 54457025 |
| 54457125 |
| 54855324 |
| 54855424 |
| 406052710 |
| 406054034 |
| 406254001 |
| 406575501 |
| 406575523 |
| 406575562 |
| 406577101 |
| 406577123 |
| 406577162 |
| 406872510 |
| 904653061 |
| 13107000000 |
| 17478000000 |
| 31722000000 |
| 42806000000 |
| 60687000000 |
| 63739000000 |
| 66689000000 |
| 67457000000 |
| 67877000000 |
| 68084000000 |
| 68462000000 |

**Supplemental Table 3.** Opioid treatment programs as reported to the Drug Enforcement Administration’s Automated Reports and Consolidated Orders System per one million persons.

| State | 2010 | 2015 | 2020 | 2021 |
| --- | --- | --- | --- | --- |
| 1. Rhode Island | 16.1 | 16.1 | 21.0 | 24.6 |
| 2. Delaware | 6.7 | 10.6 | 14.1 | 17.9 |
| 3. Maryland | 9.9 | 11.4 | 14.4 | 14.3 |
| 4. Connecticut | 10.1 | 10.3 | 11.9 | 14.1 |
| 5. Massachusetts | 7.9 | 9.1 | 12.1 | 13.7 |
| 6. Vermont | 11.2 | 14.4 | 12.4 | 12.4 |
| 7. New Mexico | 4.8 | 6.7 | 11.3 | 9.5 |
| 8. Arizona | 5.0 | 5.7 | 9.2 | 9.3 |
| 9. DC | 8.3 | 5.9 | 7.3 | 9.0 |
| 10. Alaska | 2.8 | 5.4 | 5.5 | 8.2 |
| 11. Maine | 6.8 | 9.0 | 7.3 | 7.3 |
| 12. Illinois | 5.1 | 5.6 | 7.2 | 7.2 |
| 13. Pennsylvania | 5.4 | 5.8 | 6.9 | 7.0 |
| 14. Georgia | 4.7 | 6.3 | 7.0 | 6.7 |
| 15. Kentucky | 3.0 | 3.4 | 6.2 | 6.4 |
| 16. North Carolina | 3.8 | 5.1 | 7.9 | 6.4 |
| 17. Iowa | 1.3 | 2.2 | 5.6 | 6.0 |
| 18. New York | 5.7 | 5.3 | 6.0 | 5.8 |
| 19. New Hampshire | 4.6 | 6.0 | 7.3 | 5.8 |
| 20. Ohio | 1.5 | 1.8 | 4.8 | 5.5 |
| 21. Oregon | 3.4 | 3.7 | 5.2 | 5.4 |
| 22. Nevada | 4.4 | 4.5 | 5.2 | 5.4 |
| 23. Utah | 3.6 | 4.4 | 5.5 | 5.1 |
| 24. West Virginia | 4.9 | 4.9 | 5.0 | 5.0 |
| 25. Oklahoma | 3.7 | 3.6 | 5.3 | 5.0 |
| 26. Colorado | 2.2 | 2.6 | 4.9 | 5.0 |
| 27. Michigan | 3.3 | 4.3 | 4.5 | 4.8 |
| 28. New Jersey | 0.11 | 3.7 | 4.7 | 4.7 |
| 29. Virginia | 2.1 | 3.6 | 4.5 | 4.7 |
| 30. South Carolina | 1.9 | 3.7 | 4.7 | 4.6 |
| 31. California | 3.8 | 3.8 | 4.0 | 4.2 |
| 32. North Dakota | 0.0 | 0.0 | 3.8 | 3.9 |
| 33. Wisconsin | 2.3 | 3.0 | 3.7 | 3.7 |
| 34. Montana | 2.0 | 2.9 | 3.7 | 3.6 |
| 35. Indiana | 2.2 | 2.1 | 3.4 | 3.5 |
| 36. Washington | 3.0 | 3.0 | 3.9 | 3.5 |
| 37. Alabama | 2.9 | 4.5 | 4.6 | 3.2 |
| 38. Texas | 3.3 | 3.2 | 3.3 | 3.1 |
| 39. Kansas | 2.5 | 3.1 | 3.1 | 3.1 |
| 40. Minnesota | 1.5 | 3.3 | 2.8 | 3.0 |
| 41. Missouri | 1.7 | 2.5 | 2.6 | 2.9 |
| 42. Tennessee | 1.9 | 1.8 | 2.3 | 2.9 |
| 43. Louisiana | 1.8 | 2.1 | 2.2 | 2.8 |
| 44. Hawaii | 2.9 | 2.8 | 2.8 | 2.8 |
| 45. Florida | 1.3 | 2.6 | 2.9 | 2.5 |
| 46. Idaho | 1.9 | 1.8 | 2.2 | 2.1 |
| 47. Arkansas | 1.0 | 2.0 | 2.0 | 2.0 |
| 48. South Dakota | 0.0 | 1.2 | 1.1 | 1.1 |
| 49. Nebraska | 1.6 | 1.6 | 1.5 | 1.0 |
| 50. Mississippi | 0.3 | 0.3 | 1.7 | 1.0 |
| 51. Wyoming | 0.0 | 0.0 | 0.0 | 0.0 |
| **Mean** | **3.8** | **4.6** | **5.7** | **5.9** |
| **SEM** | **0.4** | **0.5** | **0.6** | **0.6** |

## Acknowledgments

Thank you to Iris Johnston for providing access to journal articles, and Olapeju Simoyan, MD for providing feedback.

